# An intraoperative methylene blue test can guide patient selection for totally tubeless PCNL

**DOI:** 10.1101/2025.10.02.25336921

**Authors:** Jean Lee, Natalie Meyer, Dyandra Parikesit, Brian Dick, Jorge Mena, Tad Manalo, Hunter Flores, Ryan Ferreira, Mustafa Saeed, Melissa Wong, Marshall Stoller, Thomas Chi, Wilson Sui, Heiko Yang

## Abstract

**Introduction and Objective:** Totally tubeless percutaneous nephrolithotomy (PCNL) has been demonstrated to be feasible and safe, particularly for mini-PCNL. Having an objective test to guide patient selection could improve confidence in this practice and facilitate broader adoption. The purpose of this study was to evaluate the utility of an intraoperative methylene blue test to determine candidacy for totally tubeless PCNL.

**Methods:** Adult patients undergoing PCNL at four institutions were included in this study. Patients with prior reconstructive surgery, chronic pain, or requiring ureteral dilation were excluded. After stone clearance, 10 mL methylene blue (MB) dye was injected into the percutaneous access tract. A positive test (“pass”) was defined as visualization of blue dye in the bladder catheter within two minutes of injection. In some standard PCNL cases, totally tubeless management was simulated by placement of a capped nephrostomy tube. Post-operative complications within 7 days were evaluated as the primary outcome.

**Results:** The intraoperative MB test was performed in 91 PCNL cases: 60 mini-PCNLs (16 Fr access) and 31 standard PCNLs (24+ Fr access). Mini-PCNL cases were more likely to pass the MB test compared to standard PCNL cases (75% vs 38.7%, p = 0.001). Of the 45 mini-PCNL subjects who passed, 40 were left totally tubeless, resulting in two minor complications (urinary retention, obstructing ureteral stone fragment). In the standard PCNL cohort, 12 passed the MB test. Five were left totally tubeless and 3 simulated totally tubeless management with capped nephrostomy tubes; one subject had a transient postoperative creatinine elevation, and one required uncapping for steinstrasse. There were no other obstructive or infectious complications, and no surgical reintervention was required.

**Conclusions:** An intraoperative methylene blue test is a simple test with high positive-predictive value to determine candidacy for totally tubeless PCNL in real time for both mini and standard PCNLs.

## INTRODUCTION

Percutaneous nephrolithotomy (PCNL) remains the primary surgical approach for large and complex renal calculi, offering high stone-free rates and proven long-term effectiveness when less invasive options are insufficient.^1,2^ Traditionally, PCNL concludes with placement of a nephrostomy tube or ureteral stent to ensure post-operative renal drainage, tamponade bleeding, and provide access for potential second-look procedures. However, these tubes are associated with significant side effects, including increased post-operative pain, prolonged hospitalization, and elevated risk for infection.^3–5^

To address the limitations of this practice, totally tubeless PCNL has emerged as an appealing alternative. This approach eliminates both the nephrostomy tube and ureteral stent to enhance patient comfort, reduce postoperative pain, and promote faster recovery. Randomized controlled trials and meta-analyses have demonstrated that totally tubeless PCNL significantly lowers analgesic requirements, shortens hospital stay, and achieves comparable stone-free and complication rates compared to conventional approaches.^1,6–8^ In parallel, mini-PCNL, which uses smaller tract sizes under 22 Fr, has emerged as a natural complement to this strategy due to its favorable safety profile and comparable efficacy.^5,9–12^ As a result, it is increasingly combined with totally tubeless approaches.

Despite these promising results, totally tubeless PCNL has not been widely adopted in clinical practice due to concerns regarding postoperative obstruction, urine extravasation, and the lack of objective criteria to guide intraoperative decision-making.^3,6,13^ Many urologists remain hesitant to omit drainage tubes without a clear indication that the urinary system is unobstructed and adequately draining.

Recent reviews and institutional series have demonstrated that totally tubeless PCNL can be performed safely, including in patients with complex stone disease, when careful patient selection is employed. These studies report comparable complication rates and favorable postoperative outcomes, supporting the feasibility of omitting routine drainage in appropriately selected patients.^14^ However, patient selection in these series is largely based on surgeon judgment and predefined criteria, and there remains no standardized, real-time intraoperative method to functionally assess antegrade urinary drainage at the conclusion of PCNL.

To address this clinical gap, we investigated the use of an intraoperative methylene blue (MB) test as a simple, real-time method to assess antegrade urinary flow. By injecting MB dye through the percutaneous tract and observing for rapid dye efflux via the bladder, surgeons can obtain immediate feedback about ureteral patency and drainage.^15^ In this study, we evaluated the utility of an intraoperative MB test to determine candidacy for totally tubeless PCNL. We hypothesized that rapid antegrade passage of MB dye indicates that postoperative renal drainage is not needed and that these patients can be managed safely without nephrostomy tube placement or ureteral stenting.

## METHODS

This was a multi-institutional study of adult (≥18 years) patients undergoing percutaneous nephrolithotomy at four institutions (University of Colorado, University of California San Francisco, University of Michigan, and Universitas Indonesia Hospital). A total of 91 patients were included: University of Colorado (N=49), University of California San Francisco (N=36), University of Michigan (N=2), and Universitas Indonesia Hospital (N=4). Subjects were enrolled consecutively at the University of Colorado over a 9-month period in 2025; at all other institutions, subjects were enrolled nonconsecutively. Institutional ethical approval was obtained to review their data. Demographic data were extracted from the electronic medical record. Stone size was quantified on manual review of imaging and reported as cumulative diameter. All preoperative urine cultures were treated according to American Urological Association guidelines.

Mini-PCNL cases were performed using a 16 Fr x 13 cm or 15 cm ClearPetra access sheath (Well Lead) and a Holmium laser fiber. Standard PCNL cases were performed using a 24 Fr X-force dilator (Becton Dickinson) and either a Trilogy pneumatic and ultrasonic lithotriptor (Boston Scientific) or a Shockpulse ultrasonic lithotripter (Olympus). All renal access was obtained via ultrasonic and/or fluoroscopic guidance per surgeon preference. Retrograde ureteroscopy was performed at the discretion of the surgeon. Ureteroscopy was not standardized as part of the study protocol and was performed for stone clearance or diagnostic evaluation based on intraoperative judgment.

The MB test was performed at the conclusion of stone treatment (video in supplement). Exclusion criteria for performing the test included history of urologic reconstructive surgery, chronic pain, and cases requiring a ureteral access sheath. Prior to injecting MB, all instruments and catheters were removed from the ureter, and a Foley catheter was placed in the bladder. For mini-PCNL cases, the inner stylet of the Clear Petra sheath was replaced, and 10-15 mL diluted MB (~1:10 methylene blue:saline) was injected directly into the renal pelvis (video). For standard PCNL cases, MB was injected through the access sheath via a catheter-tipped syringe. A timer was set with an arbitrary cut-off of 2 minutes. This time threshold was selected pragmatically to allow sufficient opportunity for ureteral drainage while maintaining intraoperative feasibility. Injection pressure was not measured or controlled. The MB test was performed using a standardized protocol across participating institutions, including dye concentration, injection volume, and the two-minute observation period. A successful test (“pass”) was defined as visualization of MB within 2 minutes; otherwise, the test was deemed unsuccessful (“fail”) (Fig. 1).

**Figure 1.**
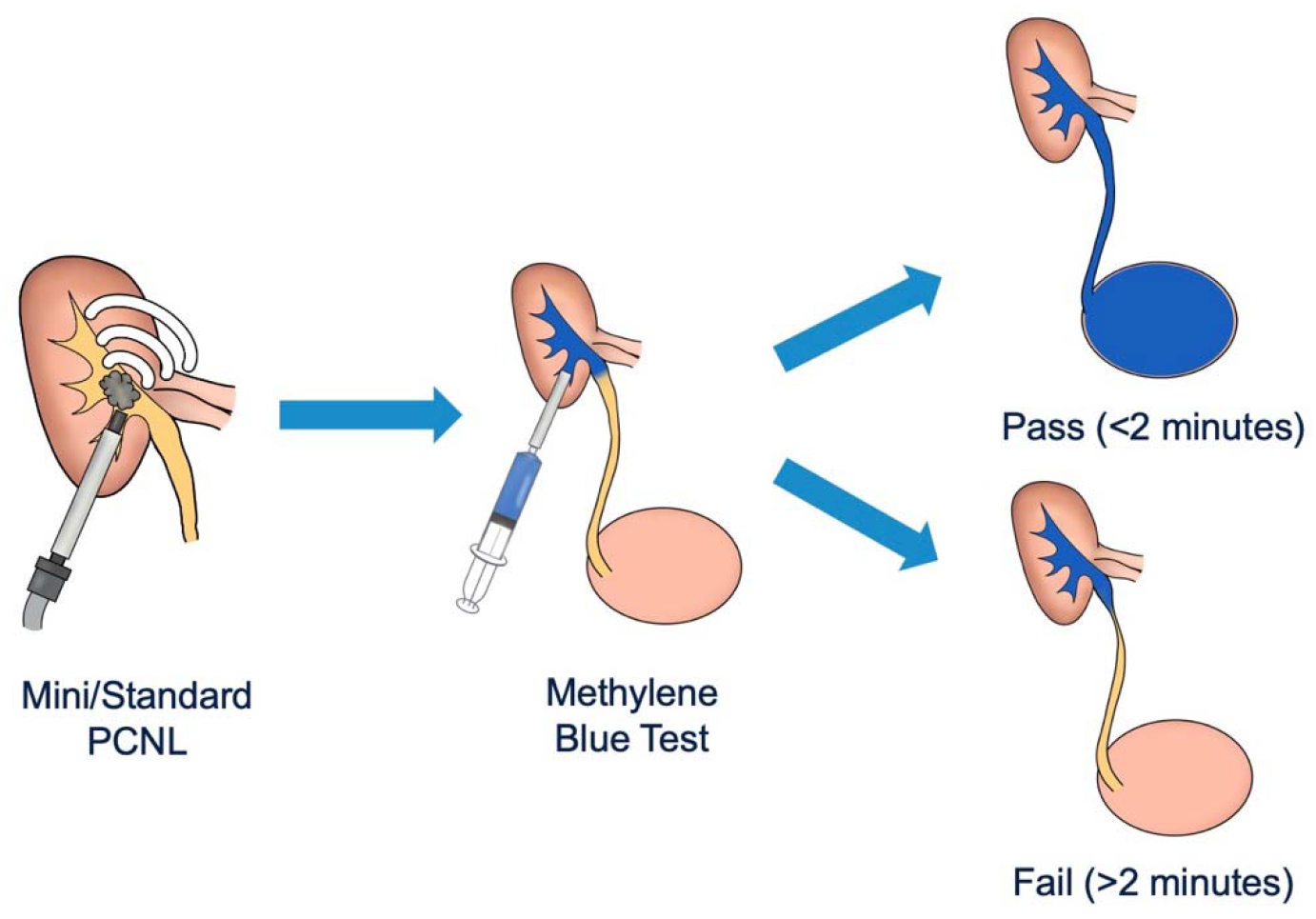
Schematic of the intraoperative methylene blue test after PCNL. Visualization of dye efflux into the bladder within 2 minutes indicates a pass and suitability for tubeless management. Absence of efflux beyond 2 minutes indicates failure requiring urinary drainage.

For mini-PCNL, a “pass” resulted in totally tubeless management, i.e. no ureteral stent or nephrostomy tube was placed, provided the following criteria were met: 1) the kidney was visually stone free on renoscopy, 2) there was no concern for ongoing hemorrhage, and 3) there was no injury to the kidney or ureter necessitating a drainage tube, e.g. mucosal perforation or ureteral damage from an impacted stone. For standard PCNL, a “pass” resulted in a simulated totally tubeless management: a nephrostomy tube was placed but was capped. As the evidence for totally tubeless management after standard PCNL is not as strong compared to mini-PCNL, this was to allow for rapid renal drainage if obstructive signs or symptoms occurred postoperatively. In both mini and standard PCNL cases, “fail” resulted in placement of a ureteral stent or nephrostomy tube per surgeon preference.

Clinical outcomes were assessed for 7 days after surgery via manual electronic medical record review. The primary outcome was complications associated with obstruction (urinary tract infection, transient fever, flank pain, nausea/vomiting, hydronephrosis on imaging, need for surgical re-intervention). Statistical analyses included Fisher’s exact test to compare relative pass rate of mini versus standard PCNL. Descriptive statistics were used to describe the primary outcomes.

## RESULTS

A total of 91 subjects underwent percutaneous nephrolithotomy (PCNL) during the study period, consisting of 60 mini-PCNL and 31 standard PCNL procedures (Table 1). Baseline demographics and clinical characteristics were similar between the two cohorts, including body mass index (BMI) (27.9 ± 6.4 vs. 26.4 ± 7.0) and American Society of Anesthesiologists (ASA) scores. Most subjects in both groups were classified as ASA 2 or 3.

**Table 1.** Baseline demographic and clinical characteristics of subjects undergoing mini-PCNL and standard PCNL. Variables include age, sex, BMI, ASA score, laterality, stone burden, operative time, and estimated blood loss.

|  | Mini (N=60) |  | Standard (N=31) |  |
| --- | --- | --- | --- | --- |
|  | Mean / No. | SD / % | Mean / No. | SD / % |
| Age | 53.6 | 16.2 | 51.9 | 16.9 |
| Gender |  |  |  |  |
| Male | 33 | 55% | 15 | 48% |
| Female | 27 | 45% | 16 | 52% |
| Race |  |  |  |  |
| White | 42 | 70% | 14 | 45% |
| Black | 4 | 7% | 6 | 19% |
| Asian | 1 | 2% | 7 | 23% |
| Other | 13 | 21% | 4 | 13% |
| BMI | 27.9 | 6.4 | 26.4 | 7.0 |
| ASA Score |  |  |  |  |
| 1 | 2 | 3% | 4 | 13% |
| 2 | 24 | 40% | 15 | 48% |
| 3 | 33 | 55% | 11 | 36% |
| 4 | 1 | 2% | 1 | 3% |
| Laterality |  |  |  |  |
| Left | 35 | 58% | 15 | 48% |
| Right | 25 | 42% | 16 | 52% |
| Stone Burden (cm) | 2.8 | 1.5 | 3.4 | 1.3 |
| Operative Time (min) | 100 | 43 | 114 | 66 |
| EBL (mL) | 56 | 10-63 | 66 | 10-500 |

|  | Mini (N=60) |  | Standard (N=31) |  |
| --- | --- | --- | --- | --- |
|  | Mean /No. | SD / % | Mean /No. | SD / % |
| *Age* | 53.6 | 16.2 | 51.9 | 16.9 |
| *Gender* |  |  |  |  |
| *Male* | 33 | 55% | 15 | 48% |
| *Female* | 27 | 45% | 16 | 52% |
| *Race* |  |  |  |  |
| *White* | 42 | 70% | 14 | 45% |
| *Black* | 4 | 7% | 6 | 19% |
| *Asian* | 1 | 2% | 7 | 23% |
| *Other* | 13 | 21% | 4 | 13% |
| *BMI* | 27.9 | 6.4 | 26.4 | 7.0 |
| *ASA Score* |  |  |  |  |
| 1 | 2 | 3% | 4 | 13% |
| 2 | 24 | 40% | 15 | 48% |
| 3 | 33 | 55% | 11 | 36% |
| 4 | 1 | 2% | 1 | 3% |
| *Laterality* |  |  |  |  |
| *Left* | 35 | 58% | 15 | 48% |
| *Right* | 25 | 42% | 16 | 52% |
| *Stone Burden (cm)* | 2.8 | 1.5 | 3.4 | 1.3 |
| *Operative Time (min)* | 100 | 43 | 114 | 66 |
| *EBL (mL)* | 56 | 10-63 | 66 | 10-500 |

The mean stone burden was 2.8 ± 1.5 cm in the mini-PCNL group and 3.4 ± 1.3 cm in the standard PCNL group. Mean operative time was shorter in the mini-PCNL cohort (100 minutes vs. 114 minutes), and estimated blood loss (EBL) was slightly lower (56 mL vs. 66 mL).

Among the mini-PCNL cohort, 45 of 60 subjects (75%) passed the MB test. Only 12 of 31 subjects (38.7%) in the standard PCNL cohort passed the MB test (p = 0.001, two-sided Fisher’s exact test) (Fig. 2).

**Figure 2.**
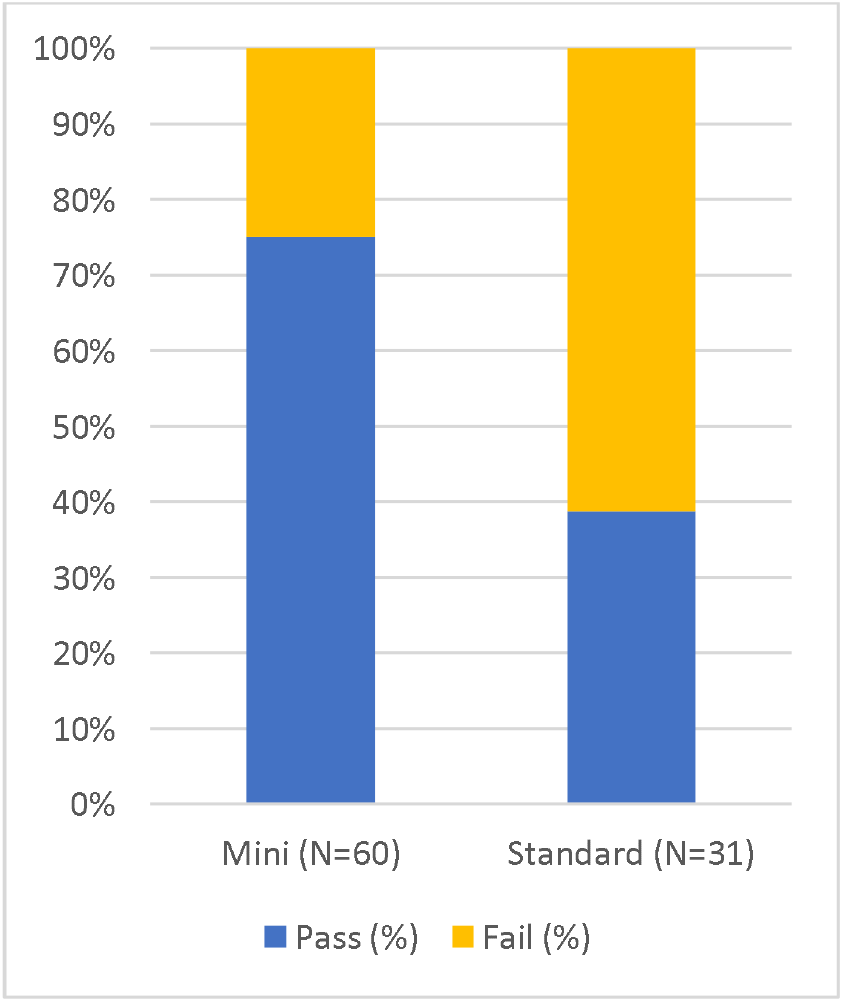
Proportion of subjects with a positive intraoperative methylene blue (MB) test in the mini-PCNL and standard PCNL cohorts. Mini-PCNL demonstrated a significantly higher MB test pass rate (*p* = 0.001, Fisher’s exact test).

Retrograde ureteroscopy was performed in a substantial proportion of cases across both cohorts. In the mini-PCNL group, ureteroscopy was performed in 31 of 45 subjects (68.9%) who passed the MB test and 12 of 15 subjects (80%) who failed the test. In the standard PCNL group, ureteroscopy was performed in 7 of 12 subjects (58.3%) who passed the MB test and 10 of 19 subjects (52.6%) who failed the test.

Of the 45 mini-PCNL subjects who passed the MB test, 40 (88.9%) were left totally tubeless postoperatively (Fig. 3). Five subjects who passed the MB test were managed with drainage tubes (nephrostomy tube or ureteral stent) at the discretion of the surgeon. Of note, 38 were discharged the same day (LOS 0 days), and none had a documented UTI within 7 days. Two postoperative complications (5%) occurred in the totally tubeless subgroup: one subject presented to the emergency department (ED) with flank pain on postoperative day (POD) 1 from a distal ureteral stone fragment that passed spontaneously. The other subject presented to the ED on POD 1 with urinary retention and acute kidney injury (AKI) that resolved with temporary bladder catheterization. Neither case required renal drainage with ureteral stenting or nephrostomy tube placement.

**Figure 3.**
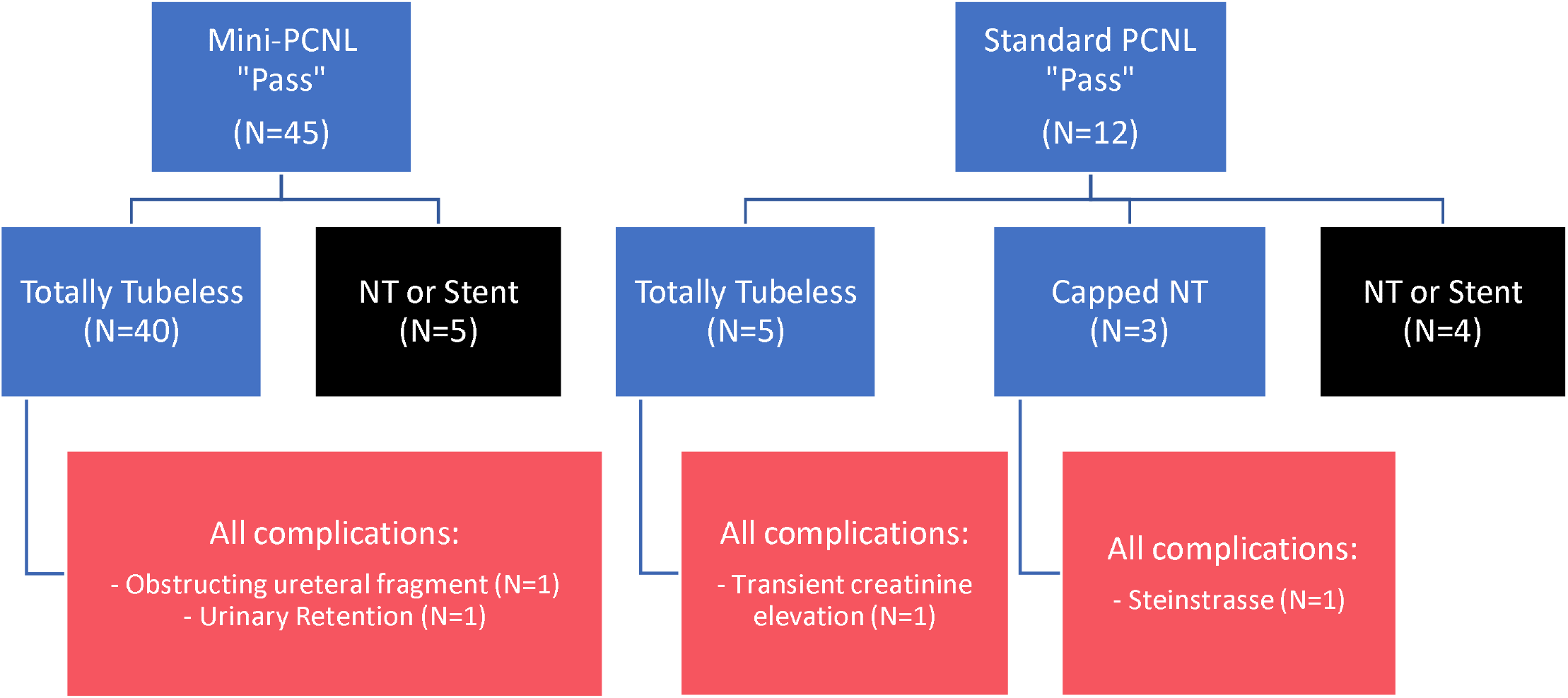
Outcomes of “passed” MB tests by surgical approach. NT = Nephrostomy tube.

In the standard PCNL cohort, 12 subjects passed the MB test. Of these, five were managed totally tubeless, with one patient developing an asymptomatic transient creatinine elevation on POD 1 that resolved spontaneously by POD2. Of the three patients who underwent simulated totally tubeless management with capped nephrostomy tubes, one required uncapping due to steinstrasse, which eventually resolved without further intervention. The remaining two subjects tolerated the capping trial without infectious or obstructive complications.

Overall, passing the MB test was associated with safe totally tubeless management, and no subjects who passed the MB test required surgical re-intervention.

## DISCUSSION

This study demonstrates that the intraoperative methylene blue (MB) test is a simple, effective method for predicting which subjects can safely undergo totally tubeless PCNL. Importantly, the MB test is intended as an intraoperative decision-support tool rather than a diagnostic assessment of fixed ureteral obstruction. By providing an objective assessment of ureteral patency in real time, the MB test addresses an important barrier to broader adoption of totally tubeless techniques.

Among 91 PCNL procedures performed at four institutions, the MB test demonstrated a high positive predictive value for safe totally tubeless management. Our findings highlight the utility of having an objective test to augment surgical judgment, which is particularly important given current PCNL practice patterns. While randomized controlled trials and meta-analyses have shown that omitting nephrostomy tubes decreases postoperative pain, shortens hospitalization, and reduces analgesic requirements without increasing major complications,^4–7^ many surgeons remain hesitant to adopt tubeless approaches due to concerns about postoperative obstruction and urine extravasation.^2,3^ The MB test addresses these concerns by providing a practical, reproducible, and cost-effective method for real-time confirmation of unobstructed urinary drainage, reducing uncertainty and supporting broader implementation of totally tubeless PCNL.^15^

Notably, despite this growing evidence base, adoption of totally tubeless PCNL remains uneven across practice settings. Recent prospective data from high-volume centers have further demonstrated that outpatient totally tubeless standard PCNL can be performed safely, with same-day discharge, and low rates of obstruction, readmission, or reintervention.^16^ These findings underscore that totally tubeless management is feasible in experienced hands. However, such approaches rely primarily on surgeon-driven selection criteria rather than an intraoperative functional assessment of urinary drainage. In this context, the reproducibility of our findings across multiple centers strengthens the evidence supporting the methylene blue test as a generalizable decision-support tool. Multi-center validation is particularly important for procedural innovations in endourology, as outcomes can be influenced by variations in patient demographics, surgeon experience, and perioperative protocols.^2,14^ Our findings therefore complement this growing body of literature by proposing a single, real-time adjunct to surgical judgment that may help facilitate broader adoption of totally tubeless PCNL across diverse practice settings and experience levels.

In both cohorts, two of the four postoperative complications were related to residual stone fragments falling into the ureter. Although these cases were managed conservatively, this finding does highlight the need for diligence in ensuring stone free status prior to proceeding with totally tubeless management. The higher pass rate among subjects undergoing mini-PCNL compared to those undergoing standard PCNL in our study supports the growing evidence that smaller access tracts and/or the use of laser instead of mechanical lithotripsy may align well with totally tubeless management. However, patient selection and case complexity for mini vs standard PCNL were likely confounding variables.^10–12^

It is also possible that concurrent ureteroscopy may impact the outcome of the MB test due to transient ureteral edema following instrumentation.^17,18^ In the present study, ureteroscopy was performed in a substantial proportion of cases across both mini and standard PCNL cohorts. In the mini-PCNL group, a higher proportion of patients who failed the MB test had undergone ureteroscopy compared to those who passed (80% vs 68.9%), suggesting a potential contribution of recent instrumentation to delayed dye transit. However, this was not statistically significant, and the relationship was not observed in the standard PCNL cohort (52.6% vs 58.3%), and the majority of patients who passed the MB test overall had also undergone ureteroscopy. These findings suggest that while ureteroscopic instrumentation may introduce variability in dye transit, the MB test remains applicable in the setting of ureteral manipulation. Importantly, any such effects would be expected to favor more cautious intraoperative decision-making.

Concerns regarding injection pressure, collecting system volume, and intrapelvic pressure merit specific discussion. Classic urodynamic and Whitaker tests are designed to diagnose fixed anatomic obstruction through pressure-based measurements. In contrast, the methylene blue test is intended as a functional screening tool to confirm antegrade urinary drainage under physiologic conditions driven by ureteral peristalsis. Prior prospective work has demonstrated that methylene blue injection, without controlling for intrapelvic pressure or individualized collecting system volume, provides predictive performance comparable to antegrade nephrostography for assessing drainage after PCNL.^15^

Several limitations of this study must be considered when interpreting the results. It is important to note that collecting system volume was not individualized in this study, as the objective of the test was confirmation of dye transit rather than generation of a target intrapelvic pressure. While extremely dilated systems could theoretically delay dye passage, such scenarios would bias toward conservative management, aligning with the MB test’s role as a safety-oriented decision aid rather than a diagnostic obstruction test. There are also differences in MB administration through the mini versus standard PCNL access sheaths. It is more difficult to prevent the dye from refluxing out of larger sheaths, so the lower passage rate in standard PCNL could be due to administration technique. Many of the subjects were discharged from the recovery room on day of surgery, which could lead to under-detection of postoperative outcomes such as transient fever or renal colic. Third, in select standard PCNL cases, a capped nephrostomy tube was used as a conservative safety measure during early adoption of the MB test. Capped nephrostomy drainage does not replicate a fully closed tubeless system, as the presence of a tube within the tract may alter local resistance to urinary extravasation and does not provide the same tissue coaptation that occurs when no tube is left in place. This practice reflected surgeon comfort at the time rather than methodological equivalence. Fourth, the negative predictive value of the MB test was not evaluated; we felt that leaving patients totally tubeless after failing the MB test was too high risk for this initial study. Finally, the study was conducted across a limited number of institutions. Future multi-center randomized trials would be beneficial to validate the accuracy and reproducibility of the MB test.

The clinical relevance of totally tubeless PCNL extends beyond its established perioperative benefits to its role in shaping future kidney stone surgery practices. In an era where enhanced recovery protocols and ambulatory urological surgery are gaining momentum, eliminating routine drainage represents a pivotal step towards same-day discharge and reduced healthcare resource utilization.^4,7^ For patients, this translates into improved comfort, a faster return to normal activity and decreased postoperative complications, all of which have a measurable impact on quality of life and satisfaction.^6,7^ For healthcare systems, fewer readmissions related to tube-associated pain, infection, or accidental dislodgement could reduce costs.^7,13^ Importantly, the safe implementation of totally tubeless PCNL relies on objective intraoperative tools like the MB test, which can standardize as well as support consistent surgical decision-making.^15^ With the global rise in stone disease, integrating these strategies into routine practice may help meet growing demand while improving safety and effectiveness.^2,8,9^

## CONCLUSIONS

An intraoperative MB test offers a simple, low-cost, and objective method to assess ureteral patency and identify candidates for totally tubeless PCNL. This technique may help reduce unnecessary drainage while maintaining patient safety, particularly in mini-PCNL cases.

## Data Availability

All data produced in the present study are available upon reasonable request to the authors.

## INSTITUTIONAL REVIEW BOARD APPROVAL

Ethics committee/IRB of University of California, San Francisco gave ethical approval for this work (IRB# 14-14533); Ethics committee/IRB of University of Colorado gave ethical approval for this work (CO-MIRB# 24-0726); Ethics committee/IRB of University of Michigan gave ethical approval for this work (IRB# HUM00278429)

## References

1. Crook TJ, Lockyer CR, Keoghane SR, Walmsley BH. A randomized controlled trial of nephrostomy placement versus tubeless percutaneous nephrolithotomy. J Urol. Aug 2008;180(2):612–4. doi:10.1016/j.juro.2008.04.020

2. de la Rosette J, Assimos D, Desai M, et al. The Clinical Research Office of the Endourological Society Percutaneous Nephrolithotomy Global Study: indications, complications, and outcomes in 5803 patients. J Endourol. Jan 2011;25(1):11–7. doi:10.1089/end.2010.0424

3. Tirtayasa PMW, Yuri P, Birowo P, Rasyid N. Safety of tubeless or totally tubeless drainage and nephrostomy tube as a drainage following percutaneous nephrolithotomy: A comprehensive review. Asian J Surg. Nov 2017;40(6):419–423. doi:10.1016/j.asjsur.2016.03.003

4. Lee JY, Jeh SU, Kim MD, et al. Intraoperative and postoperative feasibility and safety of total tubeless, tubeless, small-bore tube, and standard percutaneous nephrolithotomy: a systematic review and network meta-analysis of 16 randomized controlled trials. BMC Urol. Jun 27 2017;17(1):48. doi:10.1186/s12894-017-0239-x

5. Pietrow PK, Auge BK, Lallas CD, et al. Pain after percutaneous nephrolithotomy: impact of nephrostomy tube size. J Endourol. Aug 2003;17(6):411–4. doi:10.1089/089277903767923218

6. Xun Y, Wang Q, Hu H, et al. Tubeless versus standard percutaneous nephrolithotomy: an update meta-analysis. BMC Urol. Nov 13 2017;17(1):102. doi:10.1186/s12894-017-0295-2

7. Gauhar V, Traxer O, Garcia Rojo E, et al. Complications and outcomes of tubeless versus nephrostomy tube in percutaneous nephrolithotomy: a systematic review and meta-analysis of randomized clinical trials. Urolithiasis. Oct 2022;50(5):511–522. doi:10.1007/s00240-022-01337-y

8. Wan C, Wang D, Xiang J, et al. Comparison of postoperative outcomes of mini percutaneous nephrolithotomy and standard percutaneous nephrolithotomy: a meta-analysis. Urolithiasis. Oct 2022;50(5):523–533. doi:10.1007/s00240-022-01349-8

9. Guler A, Erbin A, Ucpinar B, Savun M, Sarilar O, Akbulut MF. Comparison of miniaturized percutaneous nephrolithotomy and standard percutaneous nephrolithotomy for the treatment of large kidney stones: a randomized prospective study. Urolithiasis. Jun 2019;47(3):289–295. doi:10.1007/s00240-018-1061-y

10. Kandemir E, Savun M, Sezer A, Erbin A, Akbulut MF, Sarilar O. Comparison of Miniaturized Percutaneous Nephrolithotomy and Standard Percutaneous Nephrolithotomy in Secondary Patients: A Randomized Prospective Study. J Endourol. Jan 2020;34(1):26–32. doi:10.1089/end.2019.0538

11. Zeng G, Cai C, Duan X, et al. Mini Percutaneous Nephrolithotomy Is a Noninferior Modality to Standard Percutaneous Nephrolithotomy for the Management of 20-40mm Renal Calculi: A Multicenter Randomized Controlled Trial. Eur Urol. Jan 2021;79(1):114–121. doi:10.1016/j.eururo.2020.09.026

12. Knoll T, Wezel F, Michel MS, Honeck P, Wendt-Nordahl G. Do patients benefit from miniaturized tubeless percutaneous nephrolithotomy? A comparative prospective study. J Endourol. Jul 2010;24(7):1075–9. doi:10.1089/end.2010.0111

13. Ganpule A, Desai M. Fate of residual stones after percutaneous nephrolithotomy: a critical analysis. J Endourol. Mar 2009;23(3):399–403. doi:10.1089/end.2008.0217

14. Tomer N, Durbhakula V, Gupta K, et al. Is Totally Tubeless Percutaneous Nephrolithotomy a Safe and Efficacious Option for Complex Stone Disease? J Clin Med. May 31 2024;13(11) doi:10.3390/jcm13113261

15. Truesdale MD, Elmer-Dewitt M, Sandri M, et al. Methylene Blue Injection as an Alternative to Antegrade Nephrostography to Assess Urinary Obstruction After Percutaneous Nephrolithotomy. J Endourol. Apr 2016;30(4):476–82. doi:10.1089/end.2015.0594

16. Gupta K, Tomer N, Connors C, et al. Is Outpatient Totally Tubeless Standard Percutaneous Nephrolithotomy Safe and Efficacious? J Endourol. Apr 2025;39(4):336–342. doi:10.1089/end.2024.0441

17. Johnson DB, Pearle MS. Complications of ureteroscopy. Urol Clin North Am. Feb 2004;31(1):157–71. doi:10.1016/S0094-0143(03)00089-2

18. Nizzardo M, Zanetti SP, Marmiroli A, et al. Transient ureteral obstruction after mini-percutaneous nephrolithotomy is associated with stone volume and location: results from a single-center, real-life study. World J Urol. Mar 13 2024;42(1):146. doi:10.1007/s00345-024-04832-6

